# A High-Performance Liquid Chromatography-Mass Spectrometry method for simultaneous determination of dolutegravir, nevirapine, efavirenz, rifampicin and rifapentine concentrations in human plasma

**DOI:** 10.1101/2025.06.08.25329229

**Authors:** Denis Omali, Moses Ocan, Hindum Lanyero, Martin Kamilo Angwe, Barbara Castelnuovo, Allan Buzibye

**Affiliations:** Department of Pharmacology and Therapeutics, College of Health Sciences, Makerere University, Kampala, Uganda; Infectious Diseases Institute, College of Health Sciences, Makerere University, Kampala, Uganda; School of Biosecurity, Biotechnology and Laboratory Sciences, College of Veterinary Medicine, Animal Resources and Biosecurity, Makerere University, Kampala, Uganda

**Keywords:** LC-MS, therapeutic drug monitoring, HIV-TB patients, Plasma, analytical method development and validation

## Abstract

Variations in clinical pharmacokinetics and genetics, and polypharmacy affect attainment of target drug concentrations in HIV-TB co-infected patients. However, scarcity of analytical methods hinders drug concentration measurement for therapeutic drug monitoring in this doubly vulnerable population in low-and-middle income countries. This study aimed to develop and validate an LC-MS method for simultaneous determination of dolutegravir, nevirapine, efavirenz, rifampicin, isoniazid and rifapentine concentrations in plasma. ThermoScientific LCQ ion trap operated by Xcalibur™ software was used. Analyte extraction from plasma was achieved by protein precipitation using 100% acetonitrile. Injection volume was 30µl for dolutegravir, efavirenz and nevirapine and 10µl for rifampicin and rifapentine. Analyte separation was achieved on Waters Atlantis dC18 column by 400µl flow rate of mobile phase under gradient elution. Transitions used were; dolutegravir (419.13→277.00), efavirenz (315.03→316.00), nevirapine (266.16→267.10), rifampicin (822.41→790.20), rifapentine (876.45→845.50), dolutegravir-d4 (423.13→280.10) and rifampicin-d4 (826.41→795.20). Calibration curves were constructed using quadratic regression with dynamic weighting of x = y, 1/x and 1/x^2^ and accepted at r^2^ ≥ 0.95 for all validation parameters. Method linearity and recovery ranged from 0.25µg/ml to 10.00µg/ml and 86.12% to 109.89% respectively for all analytes. Intra-and inter-day accuracy ranged from 88.73% to 109.67% and 93.38% to 104.30% respectively for all analytes. Both intra-and inter-day precision ranged from 2.47 RSD to 12.39 RSD and 5.34 RSD to 16.83 RSD respectively for all analytes. The method was selective and specific with no significant ghost peak identified in blank samples at expected retention times of dolutegravir, efavirenz, nevirapine, rifampicin and rifapentine. An LC-MS method for simultaneous determination of dolutegravir, efavirenz, nevirapine, rifampicin and rifapentine in human plasma was developed and validated. The method could be useful to the Uganda Ministry of Health for therapeutic drug monitoring in HIV-TB patients in health facilities in the country.

## 1.0 Introduction

In 2023, tuberculosis caused an estimated 1.25 million deaths, of which 161 000 people were co-infected with Human Immunodeficiency Virus (HIV) [1]. Despite the availability of antiretroviral and anti-tuberculosis medicines, the treatment of HIV-TB co-infection continues to challenge clinical practice. This is attributed to both individual (such as inter-individual genetic variabilities, poor medication practices, presence of other morbidities) and drug (such as drug-drug interactions, drug-related toxicities) related factors that hinder attainment of therapeutic target [2, 3], which could be minimized and controlled by therapeutic drug monitoring (TDM). Yet, despite its benefits in the developed countries [4, 5], the practice of TDM is limited in low-and middle-income countries due to limitations in affordable analytical methods for TDM and technological advancement in chromatographic platforms such as liquid chromatography (LC) with its detection systems like the mass spectrometer (MS), ultraviolet (UV) light [6].

Dolutegravir, or either of nevirapine or efavirenz are among the first-line drugs used in the management of People Living With HIV (PLWHIV) in Uganda [7]. While Rifampicin, isoniazid, ethambutol and pyrazinamide are the first-line drugs in treatment of tuberculosis [1]. Surprisingly, in the event of HIV-TB co-infection, PLWHIV are treated for both diseases concurrently. This polypharmacy precipitates drug-drug interactions, and potentiates mal-adherence due to stigma [5, 8, 9] with consequent negative treatment outcomes in this doubly vulnerable population.

Sekaggya *et al* associated negative treatment outcomes such as mortality, tuberculosis treatment failure and low isoniazid and rifampicin concentrations in HIV-TB patients in Uganda receiving either efavirenz or nevirapine [10]. Similarly, Sekaggya *et al* and other studies reported exposure to lower drug concentrations in HIV-TB patients receiving both antiretroviral therapy and anti-tuberculosis therapy [11–14]. In a systematic review by Daskapan *et al*, they discuss the potential of TDM in identification of patients at risk of subtherapeutic first-line anti-tuberculosis drug concentrations to improve treatment response and prevent toxicity [4]. These studies and related research elsewhere underscore the relevance of TDM through antiretroviral and anti-tuberculosis drug concentrations measurement in plasma.

The use of plasma over saliva, urine, hair as matrix of choice for drug concentrations monitoring is because plasma is not significantly affected by secondary organs and individual demographics like age, gender, disease state, race [15]. Similarly, preferential use of LC-MS over other chromatographic platforms is anchored in its higher specificity, sensitivity and efficiency which allows for development of simultaneous analytical methods [16, 17]. The advantage of simultaneous methods is their ability to save time and reagents and hence associated with less operational costs compared when analyzing multiple drugs compare to single methods. Available analytical methods detected either one or part of the first-line drugs used in either HIV or tuberculosis [18–25], with no method simultaneously measuring both first-line tuberculosis and the selected antiretroviral drugs in plasma. This study therefore sought to develop and validate a LC-MS method for simultaneous determination of dolutegravir, nevirapine, efavirenz, rifampicin, isoniazid and rifapentine concentrations in human plasma.

## 2.0 Materials and Methods

### 2.1 Ethical consideration and study approval

This study was approved by the School of Biomedical Sciences research ethics committee at Makerere University College of Health Sciences (Approval number: SBS-2022-164). De-identified fresh frozen plasma from Mulago hospital blood bank was used in the preparation of standard and quality control samples. Administrative approval to obtain fresh frozen plasma was granted by the Head of Clinical Laboratories of Mulago hospital. Freshly stored plasma was obtained in three batches on 14/03/2024, 08/04/2025 and 06/05/2024. The authors were therefore not able to access any information that linked the blank plasma to their respective blood donors. Administrative approval to obtain fresh frozen plasma from Mulago hospital blood bank was granted for the entire study period.

### 2.2 Instrumentation

A ThermoScientific LCQ Fleet ion trap Liquid chromatography-mass spectrometry (LC/MSn) model (Thermo Fisher Scientific Inc. 355 River Oaks Pkwy, San Jose, CA 95134) operated by Xcalibur™ software was used in this study. Sample preparation and accessory equipment used in this study included; Ohaus analytical balance Systems (Nänikon, Switzerland), ThermoScientific automatic pipettes (Waltham, USA), vortex mixer, ThermoScientific microcentrifuge (Waltham, USA).

### 2.3 Chemicals and reagents

High grade (>99.9%, suitable for HPLC) chemicals; methanol (MERCK 1060182500), acetonitrile (MERCK 1060182500) and formic acid (MERCK 1060182500) were purchased from Sigma-Aldrich (Darmstadt, Germany). Ultra-pure water was obtained from ThermoScientific Barnstead Lab Water Purification Systems (Waltham, USA). Atlantis dC18 (3.0 x 150mm, 3μm) analytical column was purchased from Swiss Laboratories (Basel, Switzerland). High purity analytical reference substances for dolutegravir > 98%, nevirapine > 95%, efavirenz = 100%, rifampicin > 95%, isoniazid = 98%, rifapentine >95%, dolutegravir-d4 > 80% and rifampicin-d4 > 95% were purchased from Toronto Research Chemicals Inc (Toronto, Canada). Blank plasma free from both antiretroviral and anti-tuberculosis drugs for preparation of the blank, zero and non-zero plasma calibrators/standards and control samples was obtained from the blood bank at the hematology unit of Mulago hospital.

### 2.4 Sample analyte extraction and chromatographic conditions

Sample analyte extraction was achieved by protein precipitation using 100% acetonitrile: plasma sample in a ratio of 3:1 for dolutegravir, efavirenz and nevirapine and 8:1 for rifampicin and rifapentine. Chromatographic separation was accomplished by an Atlantis 18C column (3.0 × 150mm, 3μm, Waters, USA) at a column temperature of 18°C - 26°C. The mobile phase consisted of eluent C (0.1% formic acid in water) and eluent D (0.1% formic acid in 100% acetonitrile). The gradient at flow rate of 400μL/min was used over the entire run time of 5.5 minutes (Table 1). The injection volume was 10μL for rifampicin and rifapentine and 30 μL for dolutegravir, efavirenz and nevirapine.

**Table 1:** Method specific mobile phase composition, gradient and flow rate.

| Time (min) | Eluent C (%) | Eluent D (%) | Flow rate (μL) |
| --- | --- | --- | --- |
| 0.00 | 40 | 60 | 400 |
| 0.50 | 40 | 60 | 400 |
| 2.00 | 5 | 95 | 400 |
| 3.00 | 5 | 95 | 400 |
| 3.10 | 40 | 60 | 400 |
| 5.50 | 40 | 60 | 400 |

### 2.5 Mass spectrometry conditions

Ionization was achieved using Heated Electrospray Ionization (HESI) with a spray voltage of 5.00 V in positive ion mode. Mass spectrometry conditions for target drugs and internal standards were optimized by delivering a single solution containing 10μg/mL isoniazid into the MS system using a syringe pump at a constant flow rate of 5μL/min. Firstly, the precursor ions were determined by Q1 full scan, and product ions were selected by product ion scan for the subsequent transition. Then, MS parameters associated with collision-induced dissociation to form transitions were optimized by Multiple Reaction Monitoring (MRM) scan and saved for subsequent method analyses (Table 2). The dwell time of each MRM transition was set at 100 ms. The source parameters were set as following: Heater temperature = 170.00°C; Capillary temperature = 275.00°C; Capillary voltage = 34.00V; Tube lens voltage = 90.00V; Ion spray voltage = 5.00 V; Sweep gas flow rate = 0 arb; Auxiliary gas flow rate = 5 arb; Sheath gas flow rate = 29 arb.

**Table 2:**
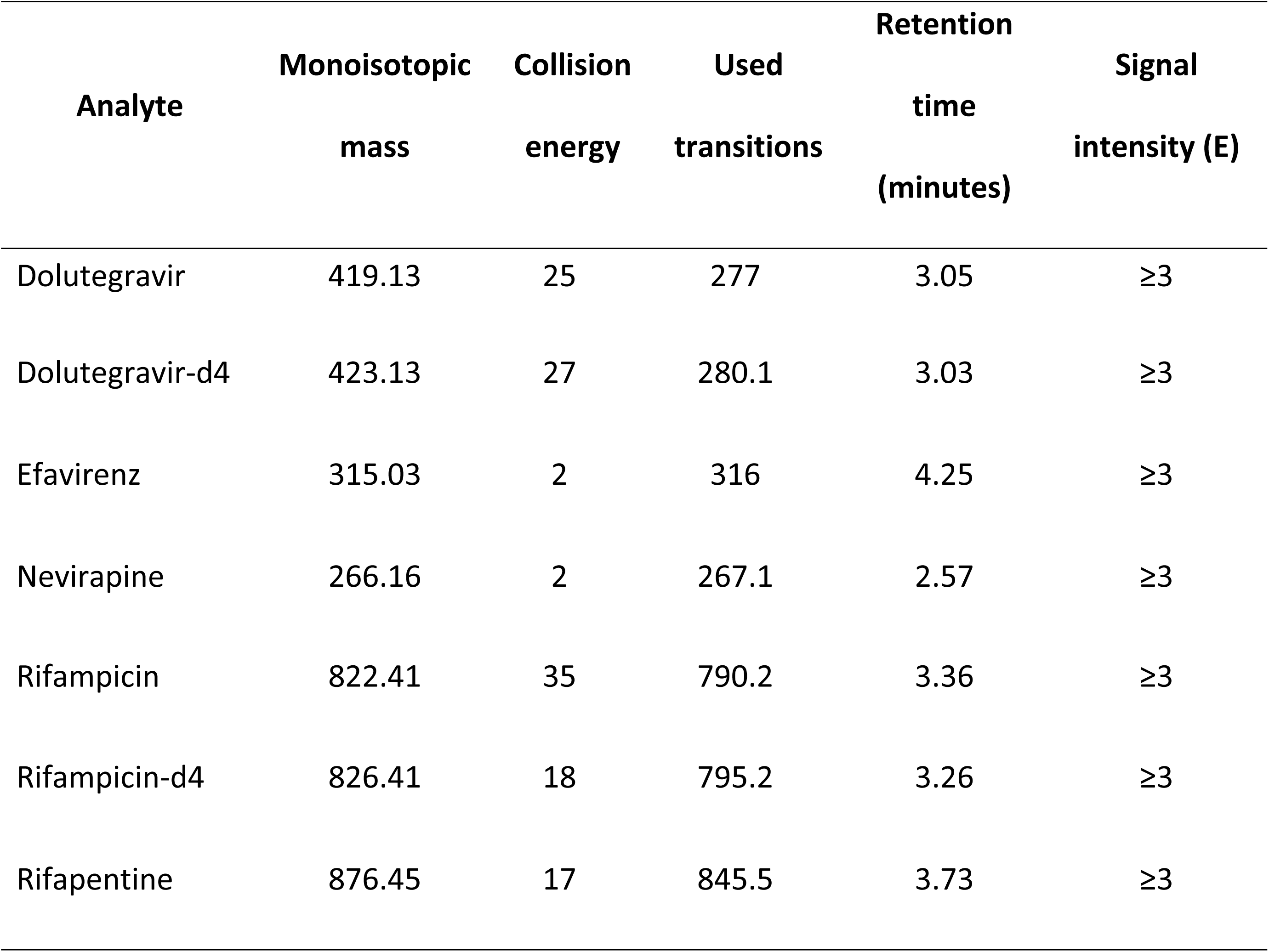
Selected analyte identification parameters for dolutegravir, nevirapine, efavirenz, rifampicin and rifapentine at optimal collision-induced dissociation energies.

| Analyte | Monoisotopic mass | Collision energy | Used transitions | Retention time (minutes) | Signal intensity (E) |
| --- | --- | --- | --- | --- | --- |
| Dolutegravir | 419.13 | 25 | 277 | 3.05 | ≥3 |
| Dolutegravir-d4 | 423.13 | 27 | 280.1 | 3.03 | ≥3 |
| Efavirenz | 315.03 | 2 | 316 | 4.25 | ≥3 |
| Nevirapine | 266.16 | 2 | 267.1 | 2.57 | ≥3 |
| Rifampicin | 822.41 | 35 | 790.2 | 3.36 | ≥3 |
| Rifampicin-d4 | 826.41 | 18 | 795.2 | 3.26 | ≥3 |
| Rifapentine | 876.45 | 17 | 845.5 | 3.73 | ≥3 |

### 2.6 Preparation of stock and working solutions

Stock solutions of 10,000 µg/ml of nevirapine, efavirenz, rifampicin and rifapentine were prepared in 100% methanol while dolutegravir and isoniazid in dimethylsulphoxide and distilled water respectively. Working solutions of 1000µg/ml (working solution 1) and 50µg/ml (working solution 2) were prepared by diluting appropriate volumes of stock solutions in a 50% water-methanol solution. Stock and working solutions for rifampicin and rifapentine were stabilized by 3% ascorbic acid solution. Internal standard-working solutions containing dolutegravir-d4 (for Dolutegravir, nevirapine and efavirenz) and rifampicin-d4 (for rifampicin and rifapentine) were separately prepared in 100% acetonitrile at a concentration of 0.00005µg/ml. The internal standard-working solutions were used as protein precipitation solutions during analyte extraction.

### 2.7 Preparation of standard curves

Standard curves were constructed starting from a blank standard (a plasma sample processed without Internal Standard-IS) followed by a zero standard (a blank plasma sample processed with IS) (Table 3). Additional six non-zero standards were included to mimic published clinical range of concentrations for, efavirenz (1-4mg/L) [26], dolutegravir (0.064-0.3mg/L) [11], nevirapine (1.6-23.0mg/L) [27] rifampicin (8-24mg/L) and rifapentine (10-30mg/L) [28]. The non-zero standard samples ranged from 0.25µg/ml 10µg/ml. Quality control samples at high, medium, and low concentrations of 8µg/ml, 4µg/ml and 0.75µg/ml respectively were prepared in the same way as calibrators.

**Table 3:** Preparation of standard and quality control samples from blank plasma and working solutions.

| STD and QC | Plasma (μL) | Working Solution (μl) | DTG, EFV, NVP, RMP, RPT (μg/ml) |
| --- | --- | --- | --- |
| BLANK | 1000 | 0 | 0.00 |
| STD1 | 995 | 5WS2 | 0.25 |
| STD2 | 990 | 10WS2 | 0.50 |
| STD3 | 950 | 50WS2 | 1.00 |
| STD4 | 995 | 5WS1 | 2.50 |
| STD5 | 975 | 25WS1 | 5.00 |
| STD6 | 950 | 50WS1 | 10.00 |
| QCL | 985 | 15WS2 | 0.75 |
| QCM | 920 | 80WS2 | 4.00 |
| QCH | 960 | 40WS1 | 8.00 |

STD; standard sample, QCL; quality control low sample, QCM; quality control medium sample, QCH; quality control high sample, WS1; working solution 1, WS2; QCL; working solution 2. DTG; dolutegravir, EFV; efavirenz, NVP; nevirapine; RMP; rifampicin and RPT; rifapentine.

### 2.8 Standard (calibration) curve/linearity

The calibration curve was constructed using standard sample concentrations with a linear range of 0.25µg/ml to 10.00µg/ml for dolutegravir, nevirapine, efavirenz, rifampicin and rifapentine. Calibration curves were constructed using both linear and quadratic regression with dynamic weighting of x = y, 1/x and 1/x2 to obtain back-calculated drug concentrations, and accepted at r2 > 0.95 for all validation parameters, with at least 75% of non-zero standard samples attaining at most 15% (and 20% for LLOQ) standard deviation of the referred value.

### 2.9 Sample processing

Two sets of standard and quality control samples were separately prepared; set one contained only rifampicin and rifapentine while set two contained dolutegravir, efavirenz and nevirapine. During sample processing for analysis, 400µL of rifampicin-d4 internal standard working solution was added to 50µL of each standard and quality control sample containing rifampicin and rifapentine. Similarly, 300µL of dolutegravir-d4 internal standard working solution was added to 100µL of each standard and quality control sample containing dolutegravir, efavirenz and nevirapine. Samples were then vortexed immediately and centrifuged for 10 minutes at 14,000 relative centrifugal force. The clear supernatant was then pipetted out and transferred into HPLC vials for analysis.

### 2.10 Methodological validation

The United States Food and Drug administration (US-FDA) guidelines for analytical method development and validation [29, 30] were followed for selectivity, precision, accuracy, linearity, calibration curve, lower limit of quantification (LLOQ), carryover, and stability.

#### 2.10.1 Accuracy and precision

Method accuracy and precision were determined by concurrently running a one-time injection of standard plasma samples together with six sets (for each level) of quality control (QC) samples of dolutegravir, efavirenz, nevirapine, rifampicin and rifapentine. The quality control samples were prepared in three levels as QC High (QCH), QC Medium (QCM) and QC Low (QCL). The observed concentrations of QCs were used to calculate accuracy and precision against the referred QC sample concentrations and expressed as relative standard deviation (RSD). Accuracy and precision were determined for concentration measurements on the same day (intra-day) and for three different days (inter-day) with six separate batches/sets of QC and LLOQ samples analyzed per QC level and LLOQ each day.

#### 2.10.2 Lower limit of quantification (LLOQ)

The LLOQ was considered as the lowest standard (standard 1) for this method, and identified by determining the ratio of signal of background noise at dolutegravir, nevirapine, efavirenz, rifampicin and rifapentine retention times in blank samples at a signal to noise ratio of 10:1 as previously described by Sonawane [31]. In this study, a value below the lower clinical therapeutic range that could be accurately measured precisely was evaluated against blank plasma samples for the effect of background noise before validation.

#### 2.10.3 Selectivity, specificity and carry-over

Method selectivity and specificity were determined to ascertain the ability of the bioanalytical method to measure and differentiate a drug in presence of other drugs or endogenous molecules with similar properties. To determine method selectivity, specificity and carry over, six different batches of plasma were used to prepare blank standard samples and analyzed in the same way as other standards. To assess method related carryover, prepared blank samples were analyzed after the upper limit (10µg/ml) of quantification standard sample to observe for any significant peak (at least 20% of LLOQ) that would appear at the retention times of dolutegravir, efavirenz, nevirapine, rifampicin and rifapentine [29, 31].

#### 2.10.4 Recovery/extraction efficiency

Method recovery evaluated the efficiency of method analyte extraction procedure. This was performed by comparing the response ratio of a selected standard concentration (0.5µg/ml) extracted from spiked plasma prior to processing to that in plasma after processing (un-extracted) for dolutegravir, nevirapine, efavirenz, rifampicin and rifapentine. The calculation below was used to determine method recovery as described by [29, 31].

#### 2.10.5 Analyte stability

##### 2.10.5.1 Long-term stability

Long-term (considering 28 days) stability of dolutegravir, nevirapine, efavirenz, rifampicin and rifapentine in plasma at-80°C was evaluated [29, 31]. Six sets of QC samples at each level were prepared and analyzed, first set in real time and then after 28 days.

##### 2.10.5.2 Short-term stability

###### 2.10.5.2.1 Freeze-thaw stability

Freeze-thaw stability of dolutegravir, nevirapine, efavirenz, rifampicin and rifapentine in plasma was evaluated by preparing and analyzing six separate batches/sets of QC and LLOQ samples at each of the three levels before storage for at least 12 hours at-80°C freezer. The QC samples were thawed at room temperature (18°C to 26°C), processed and analyzed together with freshly prepared standard plasma samples. The same QC samples were re-frozen, re-thawed and analyzed to achieve at least three freeze-thaw cycles described by [29, 31].

###### 2.10.5.2.2 Autosampler stability

The auto sampler stability of dolutegravir, nevirapine, efavirenz, rifampicin and rifapentine in plasma was evaluated by freshly preparing and analyzing six batches/sets of QC samples at each of the three levels against freshly prepared standard samples in real-time and 24 hours post initial analysis of the same QC samples as described by [31].

## 3.0 RESULTS

### 3.1 Recovery/extraction efficiency

The extraction efficiency ranged from 86.12% to 109.89% for dolutegravir, efavirenz and nevirapine, 103.65% for rifampicin and 100.48% for rifapentine (Table 4).

**Table 4:** Protein precipitation extraction efficiency for dolutegravir, efavirenz, nevirapine, rifampicin and rifapentine.

| Analyte | Recovery (%)<br>(n=6) | PASS/FAIL * |
| --- | --- | --- |
| Dolutegravir | 86.12 | Pass |
| Efavirenz | 88.88 | Pass |
| Nevirapine | 109.89 | Pass |
| Rifampicin | 103.65 | Pass |
| Rifapentine | 100.48 | Pass |

\*Pass/Fail consideration was based on 15 % extraction efficiency recommended by FDA bioanalytical guidelines (FDA, 2018; Sonawane *et al.*, 2014).

### 3.2 Lower limit of quantification (LLOQ)

The LLOQ considered and validated for this method was 0.25µg/ml for dolutegravir, efavirenz and nevirapine, rifampicin and rifapentine (Figures 1 and 2).

**Figure 1:**
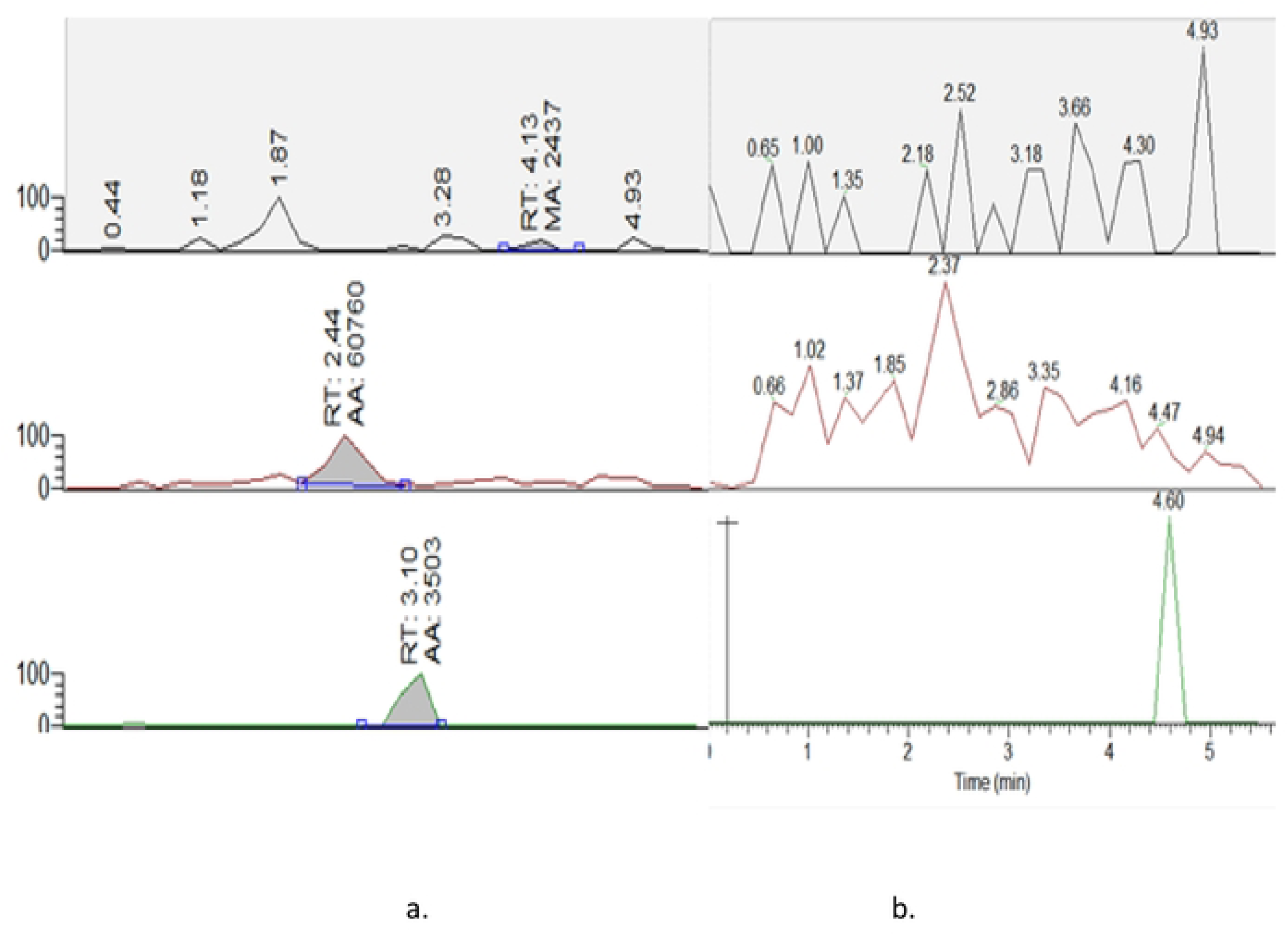
Chromatograms for a) standard-1plasma and b) blank plasma samples for efavirenz, nevirapine and dolutegravir respectively. RT-retention time, AA-analyte peak area.

**Figure 2:**
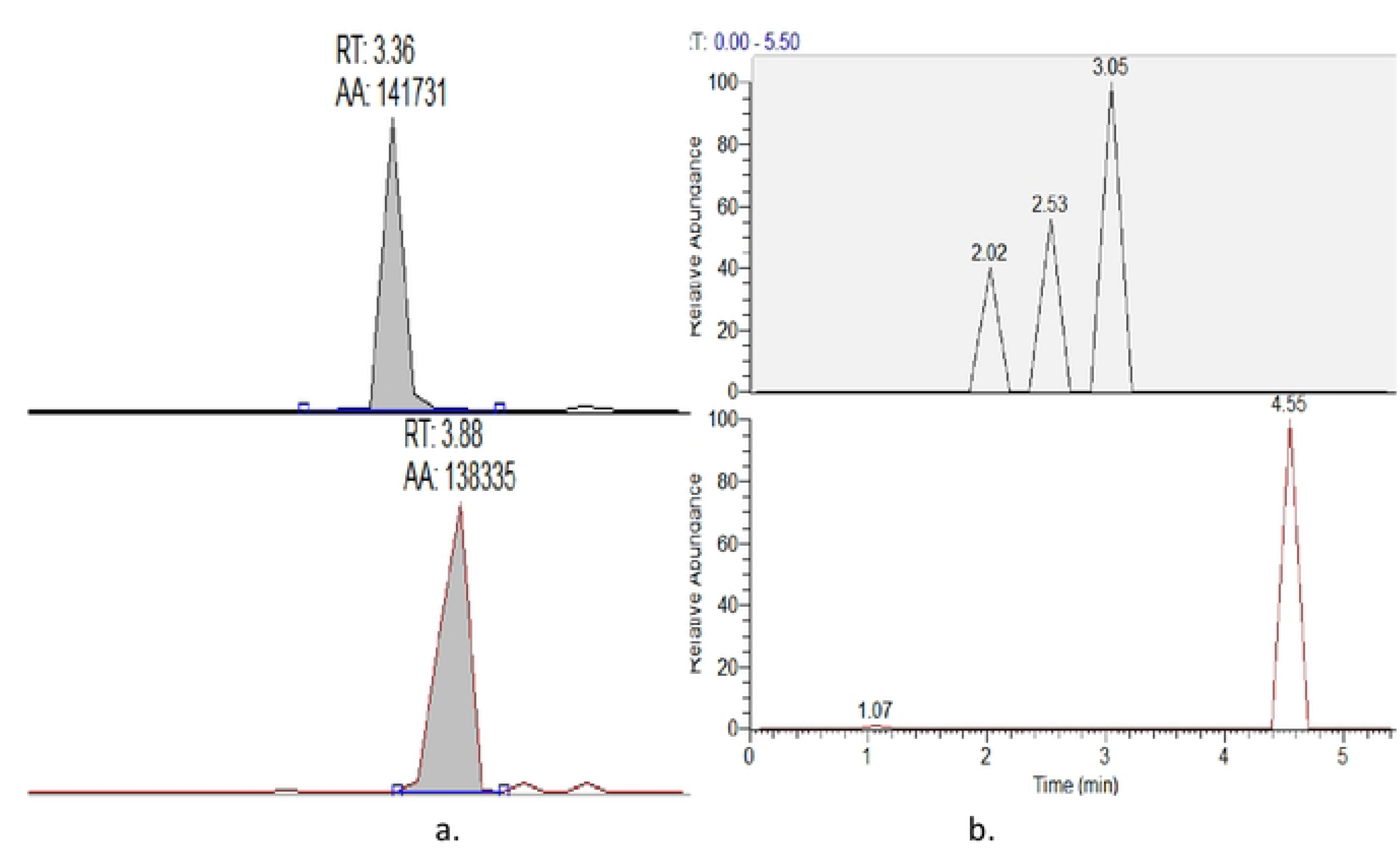
Chromatograms for a) standard-1plasma and b) blank plasma samples for rifampicin and rifapentine respectively. RT-retention time, AA-analyte peak area.

### 3.3 Selectivity, specificity and carry-over

The method was selective and specific with no carry-over above 20% of LLOQ of dolutegravir, efavirenz, nevirapine, rifampicin and rifapentine. This was indicated by absence of significant peak in six blank standard samples, and hence no interference from exogenous or endogenous compounds was observed with method specificity and selectivity as shown in blank plasma samples (Figures 1 and 2).

### 3.4 Accuracy and precision

Both intra-(2.47 to 12.39) and inter-day (5.34 to 16.83) precision were within 15% RSD of referred concentration for both QCM, QCH and 20% RSD for LLOQ and QCL for dolutegravir, efavirenz, nevirapine, rifampicin and rifapentine (Table 5).

**Table 5:**
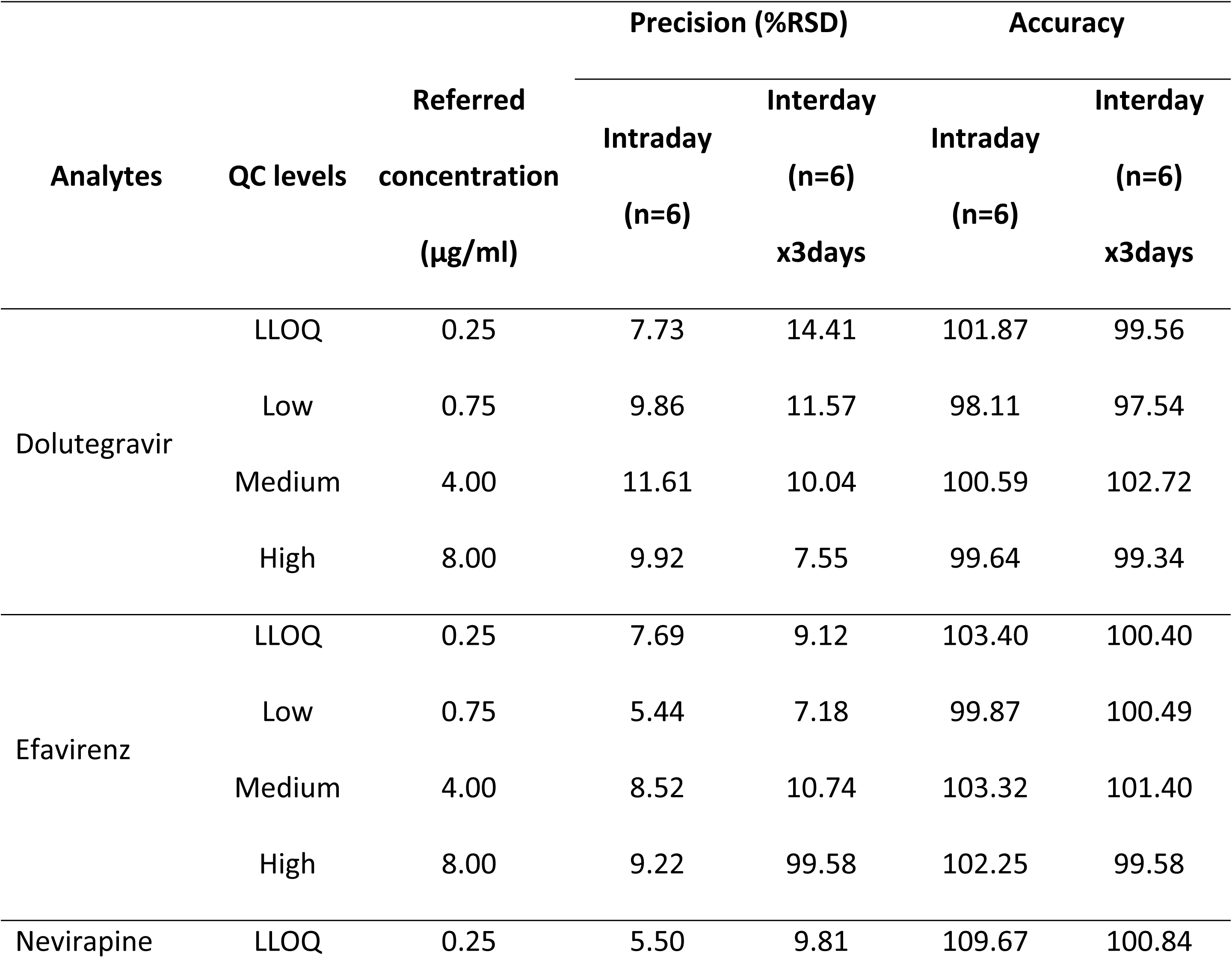
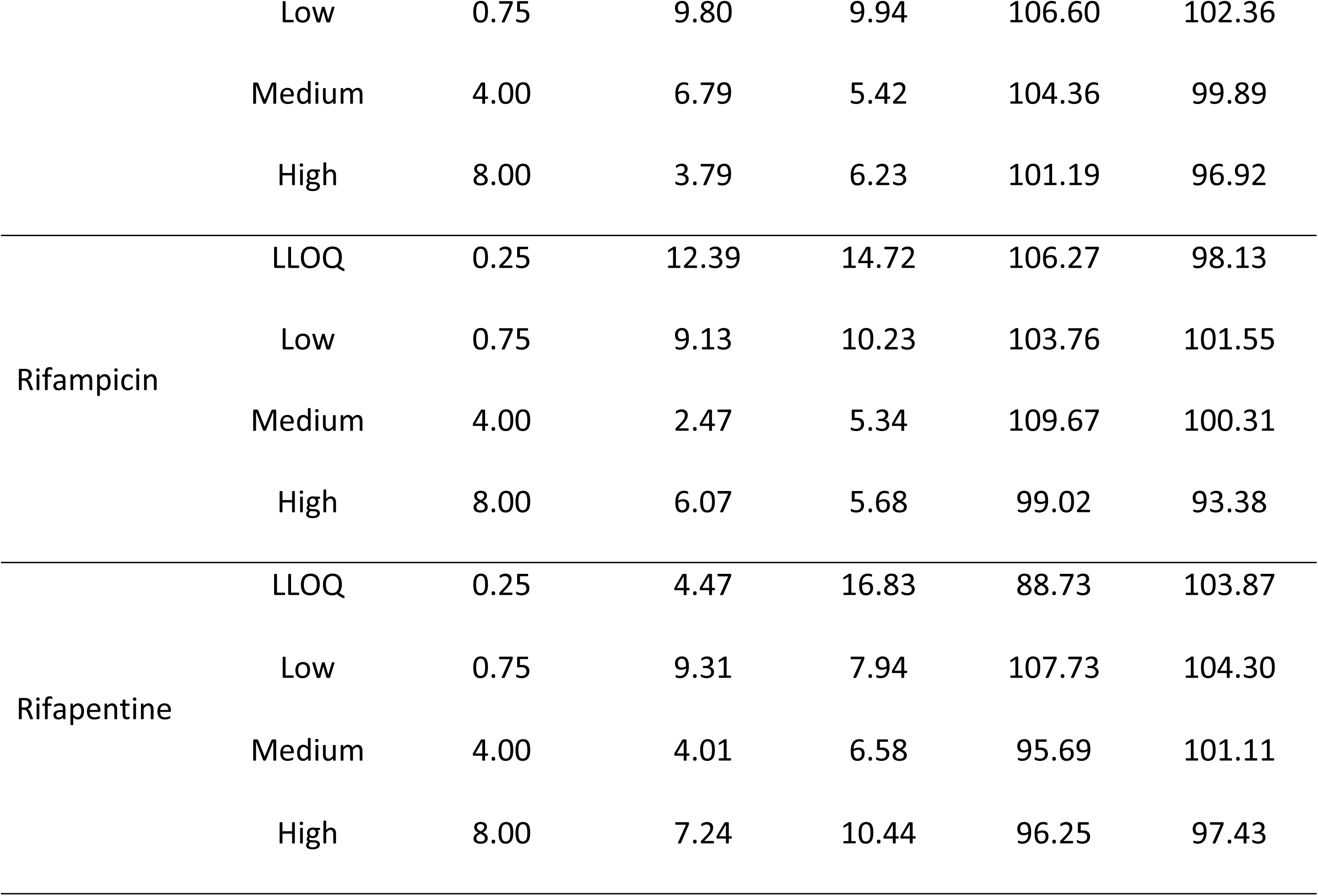
Method accuracy and precision in determining dolutegravir, efavirenz, nevirapine, rifampicin and rifapentine concentrations against referred concentrations.

Both intra-(88.73 % to 109.67 %) and inter-day (93.38% to 104.30%) accuracy were within 15 % RE of referred concentration for both QCM, QCH and 20% RE for LLOQ and QCL for dolutegravir, efavirenz, nevirapine, rifampicin and rifapentine (Table 5).

### 3.5 Analyte stability

Analyte stability was validated for long-term stability (2.47 to 12.39) and considering only freeze-thaw (5.34 to 16.83) and autosampler (3.78 to 13.30) for short-term stability. Analyte stability was within 15% CV for both QCM, QCH and 20% CV for LLOQ and QCL for dolutegravir, efavirenz, nevirapine, rifampicin and rifapentine (Table 6).

**Table 6:**
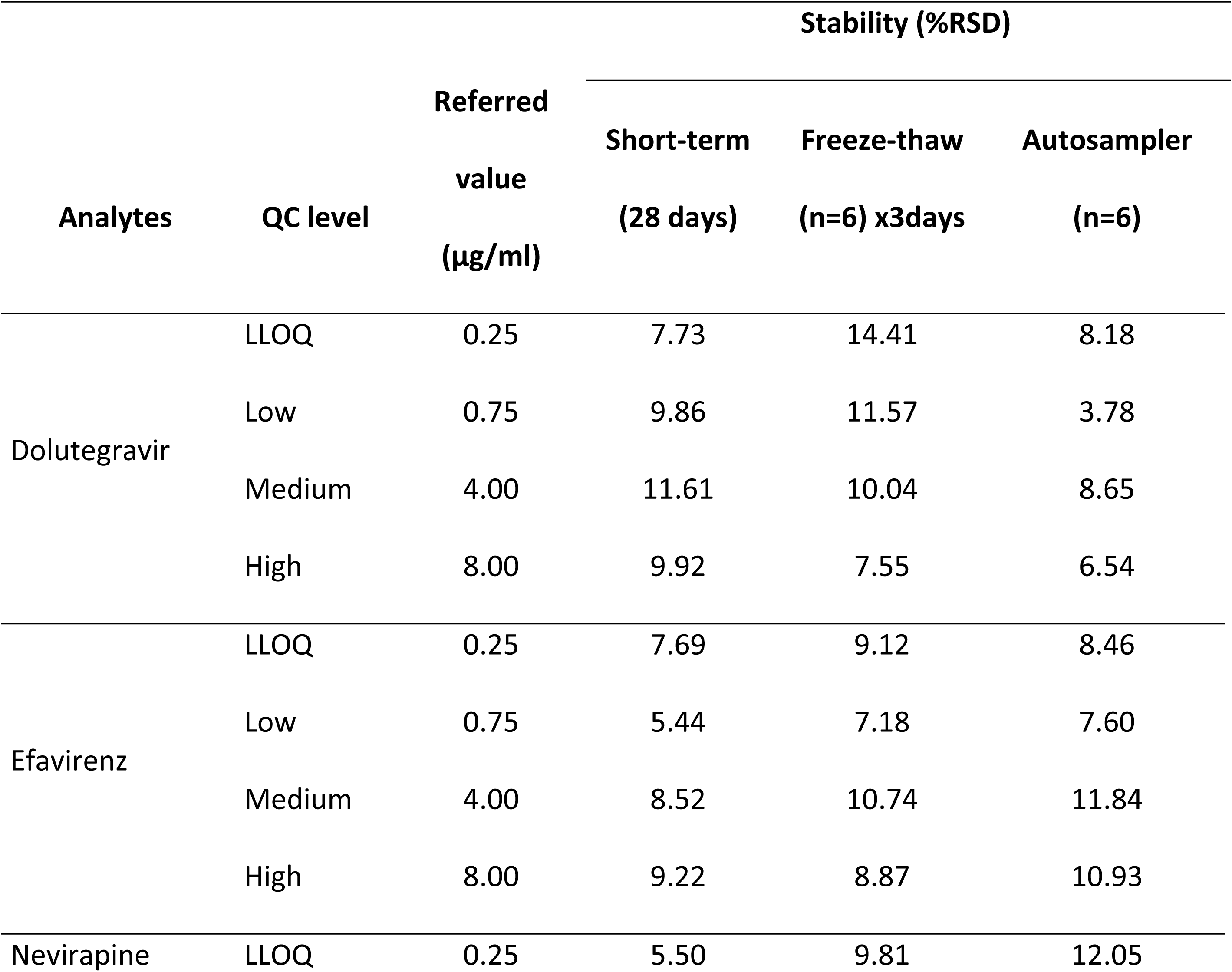
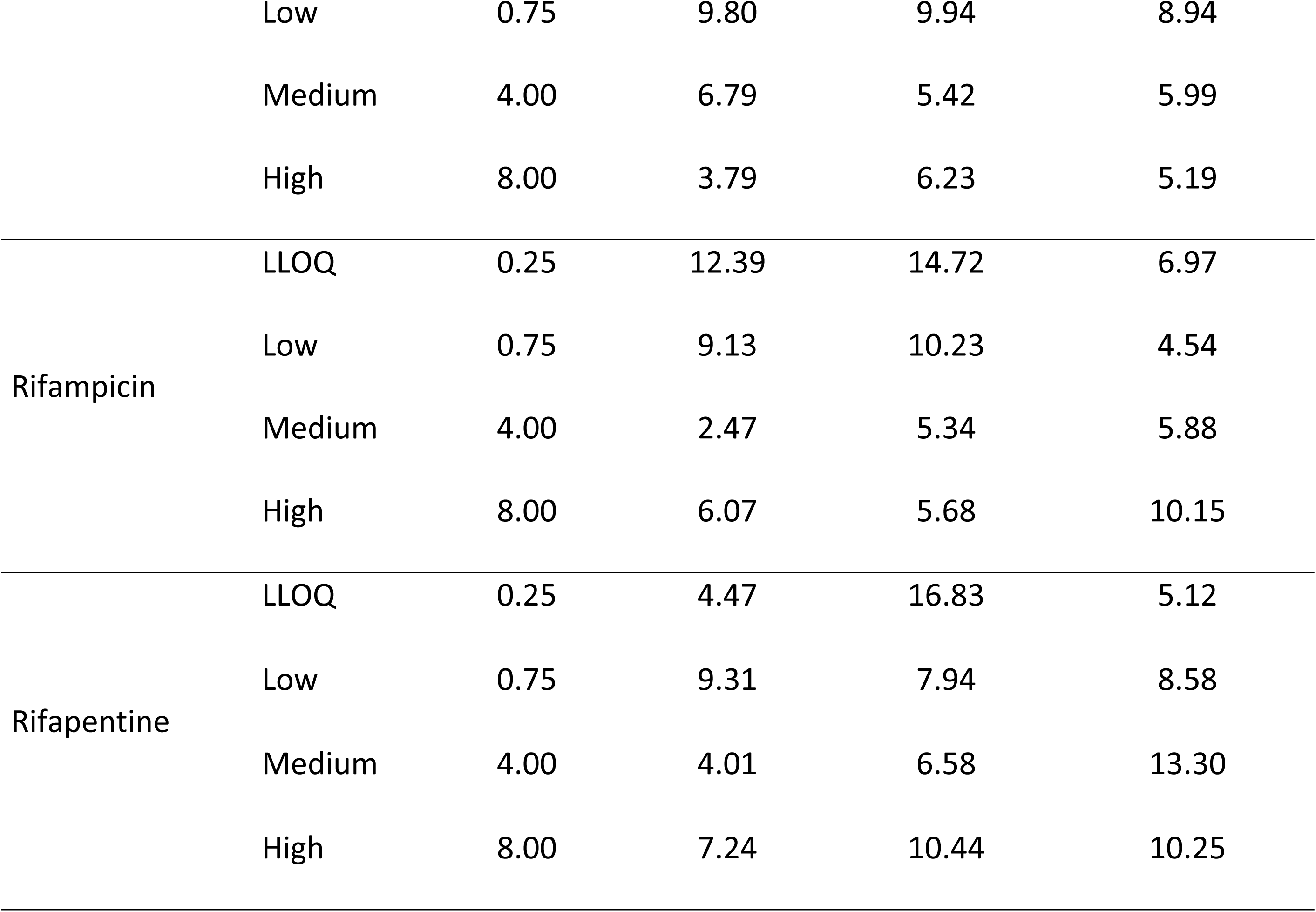
Stability of dolutegravir, efavirenz, nevirapine, rifampicin and rifapentine during method validation procedures.

## 4.0 DISCUSSION

During optimization of the study method, isoniazid was most polar than either dolutegravir, efavirenz, nevirapine, rifampicin or rifapentine due to its least LogP. This could be the most probable cause for observed irregular isoniazid peaks (not presented) with very low response and inconsistent retention times on the ^18^C column (hydrophobic and reverse phase column). Similarly, the use of 0.1 % formic acid in water (pH=2.6) and 0.1 % formic acid in 100 % acetonitrile (pH=5.8) as eluents created an acidic environment that hindered better isoniazid separation on the ^18^C column as postulated by [32]. However, unlike the polar isoniazid, other medicines; dolutegravir, efavirenz, nevirapine, rifampicin and rifapentine were best eluted at 95% organic composition of the mobile phase for 1.0 minutes. This elution was similar to that of previous studies where dolutegravir, efavirenz, nevirapine, rifampicin and rifapentine elution was achieved at high organic phase composition [18, 33, 34]. Therefore, the both the polar nature of isoniazid and method-specific acidic elution environment, acceptable isoniazid detection was not achieved for this method, prompting its elimination from the list of drug analytes. From this finding, gradient modification for future methods could enable stronger elution of both non-polar and polar analytes for inclusion of isoniazid.

The sensitivity of the current method (LLOQ = 0.25 ug/ml) was lower than that of previous studies that developed methods for determination of dolutegravir [20, 23], efavirenz [35], rifampicin [24, 36] and rifapentine [37]. However, this sensitivity was sufficient to cover the lower limit of therapeutic range of these medicines (efavirenz = 1 mg/L, nevirapine = 1.6 mg/L, rifampicin = 8 mg/L and rifapentine = 10 mg/L), except for dolutegravir = 0.064 mg/L (lower than the trough concentration, 1.123 mg/L). Similarly, the sensitivity of the current study covered the upper-limit of rifampicin MIC (0.03 to 0.25 µg/ml) [38] and rifapentine (≤ 0.12 to 0.5 µg/ml) [39] for susceptible bacilli. The method also covered dolutegravir MIC (0.21 µg/ml -< 1 µg/ml) in wild-type and resistant HIV strains [40], efavirenz therapeutic range (1.0 µg/ml to 4.0 µg/ml) [41] and nevirapine trough concentrations of 3.0 µg/ml to 4.3 µg/ml in younger children [42]. This sensitivity qualifies the method for TDM and clinical pharmacokinetics studies among PLWHIV co-infected with tuberculosis, given its accommodation of both trough and expected maximum therapeutic concentrations of the drugs.

The stability of rifapentine and rifampicin in this study was enhanced by stabilization using ascorbic acid and sample preparation in dim light similar to previous studies [33, 43]. This is because rifamycins undergo significant oxidation when exposed to air and light, leading to inaccurate concentration measurements. Because ascorbic acid is a strong antioxidant, it scavenges reactive oxygen species in plasma samples spiked with rifamycins prior to storage or sample processing [44], thereby preserving rifamycin concentrations. Freeze-thaw stability for dolutegravir was comparable to that of the method by Bennetto-Hood *et al* who used dimethylsulphoxide as in this study to prepare dolutegravir stock solution [45]. All medicines in this method were extracted from plasma by 100 % acetonitrile in which stability of drugs were shown to be preserved [18, 20, 22, 23, 33, 34, 37, 46].

However, the study was limited by lack of partial and cross validation data from other laboratories with an LC-MS in Uganda and existing inhouse assays respectively. This was mainly due to inaccessibility of other LC-MS platforms within the country, and their limited scope of analyses. Laboratories seeking to use this method should therefore optimize it on to existing analytical platforms, and perform partial validation assays prior to use in research studies or TDM.

Another limitation of the current study is that bench-top stability was not considered for validation for the analytes. This was justified by the fact that bench-top stability is not affected by method specific conditions/parameters per se, but temporal variance of analyte stay in a biological matrix at ambient temperature (18°C to 26°C). Previous studies evaluated the stability dolutegravir and efavirenz [35], nevirapine [21], rifampicin and rifapentine [25] on bench-top for 24 hours except for nevirapine that was validated for 7 hours. The studies validated the methods using only two quality control levels, low and high. The bench-top stability for both QCL and QCH respectively had a precision of 3.5 RSD and 2.5 RSD, 4.1 RSD and 2.7 RSD for dolutegravir and efavirenz; 99.79% and 101.73%, 100.06% and 102.31% accuracy. A precision of 6.8 RSD for nevirapine; and 5.2 RSD and 3.5 RSD precision for rifampicin and rifapentine respectively. These results indicate that dolutegravir, efavirenz, rifampicin and rifapentine when spiked in unextracted human plasma for 24 hours and 7 hours for nevirapine.

## 5.0 Conclusion

A sensitive HPCL-MS method was developed and validated by this study for simultaneous determination of dolutegravir, nevirapine, efavirenz, rifampicin and isoniazid concentrations in human plasma. The Uganda AIDs commission, IDI and other laboratories could consider adopting this method in future for therapeutic drug monitoring and pharmacological research.

### Glossary

1. Chromatography: The physical separation and identification of constituents of a mixture through a stationary phase (also referred to as column) by the aid of a mobile phase (also referred to as eluent/s).
2. HIV-TB co-infection: The simultaneous existence of HIV and TB infection in the same individual host/human host.
3. LogP: The partition coefficient of a solute between the organic phase and the aqueous phase.
4. LLOQ: The lowest analyte concentration that can be reliably quantified by a validated analytical method.
5. LOD: The lowest concentration of analyte that can be detected but can not be accurately quantified by a validated analytical method.
6. Minimum Inhibitory Concentration: the lowest concentration of an antimicrobial agent that prevents the visible growth of a microorganism on culture.
7. TDM: The laboratory measurement of drug concentrations in biological matrix to maximize treatment outcomes while minimizing adverse drug reactions.

## Funding

This research was supported by Fogarty International Center of the National Institutes of Health (Award #D43TW009771 “HIV and co-infections in Uganda”). The content reported here is solely the responsibility of the authors and does not necessarily represent the official views of the National Institutes of Health. Omali Denis, Castelnuovo Barbara and Buzibye Allan are partly supported by Fogarty International Center of the National Institutes of Health (Award #D43TW009771 “HIV and co-infections in Uganda”).

## CRediT authorship contribution statement

OD: Conceptualization, Investigation, Resources, Methodology, Data curation, Writing – original draft, review & editing, Supervision, Project administration, Funding acquisition for the study. OD and BA: Data curation, Validation, Formal analysis and Interpreted study data. OM, LH, AB and CB: Supervision, Methodology. OD, OM, LH, AKM, CB and BA: Writing –, review & editing for important intellectual content and writing of the final version. CB: Funding acquisition, Resources, Project administration for the study. OD provided overall coordination and technical support to the study. All authors read and approved the final manuscript version for publication.

## Declaration of competing interests

The authors declare that they have no known competing financial interests or personal relationships that could have appeared to influence the work reported in this paper.

## Data Availability

All relevant data are within the manuscript and its Supporting Information files.

## ACKNOWLEDGEMENTS

We appreciate the Capacity Building Unit of the Research Department at the Infectious Diseases Institute for the administrative support offered during the time of this research.

## Notes

### Competing Interest Statement

The authors have declared no competing interest.

### Funding Statement

Yes

### Author Declarations

This study was approved by the School of Biomedical Sciences research ethics committee at Makerere University College of Health Sciences (Approval number: SBS-2022-164).

## References

1. World Health Organisation. Global tuberculosis report 2024 2024 [Available from: https://www.who.int/teams/global-tuberculosis-programme/tb-reports/global-tuberculosis-report-2024.

2. Ashaba S, Baguma C, Tushemereirwe P, Nansera D, Maling S, Zanon BC, et al. Correlates of HIV treatment adherence self-efficacy among adolescents and young adults living with HIV in southwestern Uganda. 2024;4(9):e0003600.

3. Florence George Samizi ODP, Senga Sembuche Mulugu, Catherine Gale Gitige, Elia John Mmbaga. Rate and predictors of HIV virological failure among adults on first-line antiretroviral treatment in Dar Es Salaam, Tanzania. The Journal of Infection in Developing Countries. 2021;15(06):853–60.

4. Daskapan A, Idrus LR, Postma MJ, Wilffert B, Kosterink JG, Stienstra Y, et al. A systematic review on the effect of HIV infection on the pharmacokinetics of first-line tuberculosis drugs. 2019;58:747–66.

5. Pooranagangadevi N, Padmapriyadarsini CJFiTD. Treatment of tuberculosis and the drug interactions associated with HIV-TB co-infection treatment. 2022;3:834013.

6. Omali D, Buzibye A, Kwizera R, Byakika-Kibwika P, Namakula R, Matovu J, et al. Building clinical pharmacology laboratory capacity in low-and middle-income countries: Experience from Uganda. 2023;12(1):1956.

7. Health UMo. Consolidated guidelines on HIV prevention, diagnosis, treatment and care for key populations: World Health Organization; 2016.

8. Sundell J, Bienvenu E, Äbelö A, Ashton MJJoAC. Effect of efavirenz-based ART on the pharmacokinetics of rifampicin and its primary metabolite in patients coinfected with TB and HIV. 2021.

9. Sundell J, Wijk M, Bienvenu E, Äbelö A, Hoffmann K-J, Ashton MJAa, et al. Factors affecting the pharmacokinetics of pyrazinamide and its metabolites in patients coinfected with HIV and implications for individualized dosing. 2021;65(7):10.1128/aac.00046-21.

10. Sekaggya-Wiltshire C, von Braun A, Lamorde M, Ledergerber B, Buzibye A, Henning L, et al. Delayed Sputum Culture Conversion in Tuberculosis–Human Immunodeficiency Virus– Coinfected Patients With Low Isoniazid and Rifampicin Concentrations. Clinical Infectious Diseases. 2018;67(5):708–16.

11. Kengo A, Nabisere R, Gausi K, Musaazi J, Buzibye A, Omali D, et al. Dolutegravir pharmacokinetics in Ugandan patients with TB and HIV receiving standard-versus high-dose rifampicin. Antimicrobial agents and chemotherapy. 2023;67(11):e0043023.

12. Sekaggya-Wiltshire C, Nabisere R, Musaazi J, Otaalo B, Aber F, Alinaitwe L, et al. Decreased dolutegravir and efavirenz concentrations with preserved virological suppression in patients with tuberculosis and human immunodeficiency virus receiving high-dose rifampicin. 2023;76(3):e910–e9.

13. Sekaggya-Wiltshire C, Chirehwa M, Musaazi J, von Braun A, Buzibye A, Muller D, et al. Low antituberculosis drug concentrations in HIV-tuberculosis-coinfected adults with low body weight: is it time to update dosing guidelines? 2019;63(6):10.1128/aac.02174-18.

14. Kengo A, Gausi K, Nabisere R, Musaazi J, Buzibye A, Omali D, et al. Unexpectedly low drug exposures among Ugandan patients with TB and HIV receiving high-dose rifampicin. 2023;67(11):e00431–23.

15. Märtson A-G, Burch G, Ghimire S, Alffenaar J-WC, Peloquin CAJEOoDM, Toxicology. Therapeutic drug monitoring in patients with tuberculosis and concurrent medical problems. 2021;17(1):23–39.

16. Adaway JE, Keevil BGJJoCB. Therapeutic drug monitoring and LC–MS/MS. 2012;883:33-49.

17. Shipkova M, Svinarov DJCb. LC–MS/MS as a tool for TDM services: where are we? 2016;49(13-14):1009-23.

18. Malinga TH. LC-MS/MS method development and validation for simultaneous quantification of first-line HIV drugs and second-line TB drugs in rat plasma 2018.

19. Notari S, Bocedi A, Ippolito G, Narciso P, Pucillo LP, Tossini G, et al. Simultaneous determination of 16 anti-HIV drugs in human plasma by high-performance liquid chromatography. Journal of chromatography B, Analytical technologies in the biomedical and life sciences. 2006;831(1-2):258–66.

20. Rackow AR, Pandey A, Price AL, Marzinke MAJJoMS, Lab AitC. Rapid and sensitive liquid chromatographic–tandem mass spectrometric methods for the quantitation of dolutegravir in human plasma and breast milk. 2024;34:1–7.

21. Reddy S, Thomas L, Santoshkumar K, Nayak N, Mukhopadhyay A, Thangam SJJoAS, et al. A LC–MS/MS method with column coupling technique for simultaneous estimation of lamivudine, zidovudine, and nevirapine in human plasma. 2016;7:1–10.

22. Rezk NL, Tidwell RR, Kashuba AD. High-performance liquid chromatography assay for the quantification of HIV protease inhibitors and non-nucleoside reverse transcriptase inhibitors in human plasma. Journal of chromatography B, Analytical technologies in the biomedical and life sciences. 2004;805(2):241–7.

23. Rigo-Bonnin R, García-Tejada L, Mas-Bosch V, Imaz A, Tiraboschi JM, Scévola S, et al. Development and validation of equilibrium dialysis UHPLC-MS/MS measurement procedures for total and unbound concentrations of bictegravir, dolutegravir, darunavir and doravirine in human plasma. Application to patients with HIV. 2024;552:117678.

24. Sudha P, Dang RJIJoPE, Research. The Fast and Simple LC-MS/MS Method for Determination of Rifampicin in the Human Blood Plasma. 2024;58.

25. Winchester LC, Podany AT, Baldwin JS, Robbins BL, Fletcher CVJJop, analysis b. Determination of the rifamycin antibiotics rifabutin, rifampin, rifapentine and their major metabolites in human plasma via simultaneous extraction coupled with LC/MS/MS. 2015;104:55–61.

26. Bednasz CJ, Venuto CS, Ma Q, Daar ES, Sax PE, Fischl MA, et al. Efavirenz Therapeutic Range in HIV-1 Treatment-Naive Participants. Ther Drug Monit. 2017;39(6):596–603.

27. Cressey TR, Punyawudho B, Le Coeur S, Jourdain G, Saenjum C, Capparelli EV, et al. Assessment of Nevirapine Prophylactic and Therapeutic Dosing Regimens for Neonates. Journal of acquired immune deficiency syndromes (1999). 2017;75(5):554–60.

28. Aït Moussa L, El Bouazzi O, Serragui S, Soussi Tanani D, Soulaymani A, Soulaymani R. Rifampicin and isoniazid plasma concentrations in relation to adverse reactions in tuberculosis patients: a retrospective analysis. Therapeutic advances in drug safety. 2016;7(6):239–47.

29. Administration FaD. Bioanalytical Method Validation—Guidance for Industry. U.S. Department of Health and Human Services, Food and Drug Administration. Center for Drug Evaluation. 2018.

30. Health USDo, Human Services CfDE, Research, Center for Veterinary M. Bioanalytical Method Validation: Guid-ance for Industry. https://www.fdagov/downloads/Drugs/Guidance/ucm070107pdf2018.

31. Lalit V Sonawane BNP, Sharad V Usnale, Pradeepkumar V Waghmare and Laxman H Surwase. Bioanalytical method validation and its pharmaceutical application-a review. Pharm Anal Acta. 2014;5(288):2.

32. Atkins PWR, R George de Paula, Julio Wormald, Mark. Physical chemistry for the life sciences: Oxford University Press; 2023.

33. Fox DOC, R. Mallon, P. McMahon, G. Simultaneous determination of efavirenz, rifampicin and its metabolite desacetyl rifampicin levels in human plasma. Journal of pharmaceutical and biomedical analysis. 2011;56(4):785–91.

34. Han F, Li W, Jin Y, Wang F, Yuan B, Xu HJJoCS. Rapid and Sensitive LC-MS/MS Method for Simultaneous Determination of Three First-Line Oral Antituberculosis Drug in Plasma. 2021;59(5):432–8.

35. Xiaoying ZZ, YE Lingjie, WU Jinjin, YUAN Xiaoling, YU. Simultaneous Determination of Blood Concentrations of Five Antiretrovirals in Human Plasma by Ultra High Performance Liquid Chromatography-tandem Mass Spectormetry Method. J Chromatogr B Analyt Technol Biomed Life Sci. 2024;43(2):207–14.

36. van den Elsen SH, Oostenbrink LM, Heysell SK, Hira D, Touw DJ, Akkerman OW, et al. Systematic review of salivary versus blood concentrations of antituberculosis drugs and their potential for salivary therapeutic drug monitoring. 2018;40(1):17.

37. Fan X, Guo S, Zhang R, Cai Q, Lang Y, Huang J, et al. Development, Validation, and Clinical Application of an Ultra–High-Performance Liquid Chromatography Coupled With Tandem Mass Spectrometry Method for the Determination of 10 Antituberculosis Drugs in Human Serum. 2024;46(4):477–84.

38. Maitre T, Baulard A, Aubry A, Veziris N. Optimizing the use of current antituberculosis drugs to overcome drug resistance in Mycobacterium tuberculosis. Infectious diseases now. 2024;54(1):104807.

39. Vasiliauskaitė L, Bakuła Z, Vasiliauskienė E, Bakonytė D, Decewicz P, Dziurzyński M, et al. Detection of multidrug-resistance in Mycobacterium tuberculosis by phenotype-and molecular-based assays. 2024;23(1):81.

40. Li M, Oliveira Passos D, Shan Z, Smith SJ, Sun Q, Biswas A, et al. Mechanisms of HIV-1 integrase resistance to dolutegravir and potent inhibition of drug-resistant variants. 2023;9(29):eadg5953.

41. Martín AS, Gómez AI, García-Berrocal B, Figueroa SC, Sánchez MC, Calvo Hernandez MV, et al. Dose reduction of efavirenz: an observational study describing cost–effectiveness, pharmacokinetics and pharmacogenetics. 2014;15(7):997–1006.

42. Gopalan BP, Mehta K, D’Souza R R, Rajnala N, A KH, Ramachandran G, et al. Sub-therapeutic nevirapine concentration during antiretroviral treatment initiation among children living with HIV: Implications for therapeutic drug monitoring. PloS one. 2017;12(8):e0183080.

43. Le Guellec Gaudet M-LL, Sandrine Breteau, Michel. Stability of rifampin in plasma: consequences for therapeutic monitoring and pharmacokinetic studies. J Therapeutic drug monitoring. 1997;19(6):669–74.

44. Subashini R. Oral Delivery of ascorbic acid stabilized rifampicin nanoparticles for enhanced bioavailability of rifampicin: Swamy Vivekanandha College of Pharmacy, Tiruchengode; 2013.

45. Bennetto-Hood C, Tabolt G, Savina P, Acosta EPJJoCB. A sensitive HPLC–MS/MS method for the determination of dolutegravir in human plasma. 2014;945:225–32.

46. Lamorde M, Fillekes Q, Sigaloff K, Kityo C, Buzibye A, Kayiwa J, et al. Therapeutic drug monitoring of nevirapine in saliva in Uganda using high performance liquid chromatography and a low cost thin-layer chromatography technique. BMC Infect Dis. 2014;14:473.

